# Clinical Risk, Sociodemographic Factors, and SARS-CoV-2 Infection Over Time in Ontario, Canada

**DOI:** 10.1101/2021.04.28.21256052

**Authors:** Jacob A. Udell, Bahar Behrouzi, Atul Sivaswamy, Anna Chu, Laura E. Ferreira-Legere, Jiming Fang, Shaun G. Goodman, Justin A. Ezekowitz, Kevin R. Bainey, Sean van Diepen, Padma Kaul, Finlay A. McAlister, Isaac I. Bogoch, Cynthia Jackevicius, Husam Abdel-Qadir, Harindra C. Wijeysundera, Dennis T. Ko, Peter C. Austin, Douglas S. Lee

**Affiliations:** ICES, Toronto, Canada; Cardiovascular Division, Department of Medicine, Women’s College Hospital, Toronto, Canada; Peter Munk Cardiac Centre, University Health Network, Toronto, Canada; Institute of Health Policy, Management, and Evaluation, University of Toronto, Toronto, Canada; Department of Medicine, Temerty Faculty of Medicine, University of Toronto, Toronto, Canada; Division of Cardiology, St. Michael’s Hospital, Toronto, Canada; Canadian VIGOUR Centre, University of Alberta, Edmonton, Canada; Department of Medicine, Faculty of Medicine & Dentistry, University of Alberta, Edmonton, Canada; Department of Critical Care Medicine and Division of Cardiology, Department of Medicine, University of Alberta, Edmonton, Canada; Divisions of General Internal Medicine and Infectious Diseases, University Health Network, Toronto, Canada; Western University of Health Sciences, Pomona, CA; Schulich Heart Centre, Sunnybrook Health Sciences Centre, Toronto, Canada

**Keywords:** COVID-19, risk factors, disparities, race/ethnicity

## Abstract

**Background:** Sociodemographic and clinical factors are emerging as important predictors for developing severe acute respiratory syndrome coronavirus 2 (SARS-CoV-2) infection.

**Objective:** To determine whether public health interventions that culminated in a stay-at-home lockdown instituted during the first wave of the pandemic in March/April 2020 were effective at mitigating the association of any of these factors with the risk of infection.

**Design:** Population-based cohort study

**Setting:** Ontario, Canada

**Patients:** All adults that underwent testing for SARS-CoV-2 between January 1 and June 12, 2020.

**Measurements:** The outcome of interest was SARS-CoV-2 infection, determined by reverse transcription polymerase chain reaction testing. Adjusted odds ratios (ORs) were determined for sociodemographic and clinical risk factors before and after the peak of the pandemic to assess for changes in effect sizes.

**Results:** Among 578,263 community-dwelling individuals, 20,524 (3.5%) people tested positive. The association between age and SARS-CoV-2 infection risk among tested community-dwelling individuals varied over time (P-interaction <0.0001). Prior to the first-wave peak of the pandemic, the likelihood of SARS-CoV-2 infection increased progressively with age compared with individuals aged 18-45 years (P<0.0001). This association subsequently reversed, with all age groups younger than 85 years at progressively higher risk of infection (P<0.0001) after the peak. Otherwise, risk factors that persisted throughout included male sex, residing in lower income neighborhoods, residing in more racially/ethnically diverse communities, immigration to Canada, and history of hypertension and diabetes. While there was a reduction in infection rates across Ontario after mid-April, there was less impact in regions with higher degrees of racial/ethnic diversity. When considered in an additive risk model, following the initial peak of the pandemic, individuals living in the most racially/ethnically diverse communities with 2, 3, or ≥4 risk factors had ORs of 1.89, 3.07, and 4.73-fold higher for SARS-CoV-2 infection compared to lower risk individuals in their community (all P<0.0001). In contrast, in the least racially/ethnically diverse communities, there was little to no gradient in infection rates across risk strata.

**Conclusion:** After public health interventions in March/April 2020, people with multiple risk factors residing in the most racially diverse communities of Ontario continued to have the highest likelihood of SARS-CoV-2 infection while risk was mitigated for people with multiple risk factors residing in less racially/ethnically diverse communities. Further efforts are necessary to reduce the risk of SARS-CoV-2 infection among the highest risk individuals residing in these communities.

**Primary Funding Source:** Canadian Institutes of Health Research and the Ted Rogers Centre for Heart Research.

## Introduction

In late 2019, a novel coronavirus-2019 (COVID-19) emerged as a worldwide pandemic threat.^1^ By early 2020, virus transmission began to spread across North America. Transmission and cases of severe acute respiratory syndrome coronavirus 2 (SARS-CoV-2) infection developed earlier and in more rapid succession in certain cities with densely populated and marginalized populations.^2,3^ Other countries had some lead time to prepare for SARS-CoV-2 infections, including Canada, which has an overall smaller and more geographically distributed population size and lower scale of international travel that could influence the regional epidemiology of COVID-19.

Ontario is the most populated province in Canada with diversity in age, socioeconomic status, and race/ethnicity across the province’s counties, townships, and municipalities.^4^ While the presence of certain clinical characteristics, particularly a history of cardiovascular and renal disease or obesity, may increase the likelihood of SARS-CoV-2 infection,^5–13^ other data show sociodemographic characteristics, including race and socioeconomic status, may be stronger drivers of infection risk as seen with other communicable diseases.^14,15,24,25,16–23^ However, prior data may be biased if cases were selected only among individuals presenting to hospital, with limited or no controls. Furthermore, estimates of risk may be biased by the propensity for or against exposure and testing over time. Therefore, we sought to evaluate the association of sociodemographic and clinical risk factors with the likelihood of SARS-CoV-2 infection in Ontario by analyzing linked population-based health databases among individuals tested for SARS-CoV-2. We further evaluated whether public health interventions instituted during the first wave of the pandemic in March/April of 2020 (Supplemental Figure 1) were effective at mitigating both sociodemographic and clinical risk factors.

## Methods

### Study Design and Population

Ontario has a publicly funded health care system with universal access to care without user fees at the point of service. We assembled a population-based retrospective cohort of all Ontarians aged 18 years and older who were eligible for the province’s universal Ontario Health Insurance Plan (OHIP), alive as of January 1, 2020, and who underwent testing for SARS-CoV-2 up to June 12, 2020 (Supplemental Figure 2). This cohort was created at ICES, a non-profit research institute whose legal status under Ontario’s health information privacy law allows it to collect and analyze health care and demographic data, without consent, for health system evaluation and improvement. The cohort was created through linkage of multiple provincial and federal health care related databases (e.g., hospital discharge abstracts, physician claims, chronic disease registries, health survey, laboratory, and drug dispensing data), as well as the Immigration, Refugees and Citizenship Canada (IRCC) Permanent Resident database.^26^ These datasets were linked using unique, encoded identifiers and analyzed at ICES as previously described and validated.^27^ We required individuals to be eligible for health insurance one year prior to the index date of study as determined from the Ontario Registered Persons Database (RPDB), which includes basic demographic information about anyone who has ever had an Ontario health insurance number. Individuals who were not residents of Ontario on the index date were excluded. The index date for study inclusion was the date of a first SARS-CoV-2 test as recorded in the Ontario Laboratories Information System (OLIS) database, which consolidated the majority (88%) of results of COVID-19 testing in Ontario.

### Exposure Variables

Testing date was divided into calendar weeks. We obtained data on age, sex, and community-dwelling characteristics from the RPDB. Communities were categorized by regional public health units (PHU), geographic location, and size according to Statistics Canada’s Census data.^28^ Communities with less than 10,000 residents were classified as rural. Median neighborhood income was categorized by quintile according to national Census data. Individuals that immigrated to Ontario as their first place of landing in Canada between 1985 and 2017 were identified via the IRCC permanent resident database. Residents living in long-term care (LTC) facilities were identified via the Ontario Drug Benefit (ODB) database.

Clinical comorbidities were identified using previously validated case definition algorithms for Canadian administrative databases based on hospitalization and emergency department records from the Canadian Institute for Health Information Discharge Abstract Database (CIHI DAD) and National Ambulatory Care Reporting System (NACRS), respectively using International Classification of Diseases, Tenth Revision, Canada (ICD-10-CA) coding, hospital and physician procedure coding, and chronic disease diagnoses from the OHIP database (Supplemental Table 1). In addition to the number of hospitalization or emergency department episodes in the prior year, we also included the following characteristics within the previous 5 years: history of coronary artery disease (CAD; defined as a prior myocardial infarction, percutaneous or surgical coronary revascularization); hospitalization for heart failure (HF) or stroke, history of liver disease, chronic lung disease (including pneumonia, tuberculosis, asthma or chronic obstructive pulmonary disease), organ transplantation, atrial fibrillation, chronic kidney disease (CKD) or malignant cancer. Any prior history and duration of hypertension, any history of diabetes or human immunodeficiency virus (HIV) was also assessed. Frailty was defined using the Johns Hopkins’ Adjusted Clinical Groups (ACG®) Version 10 frailty indicator.^29,30^ Communities were categorized according to Ontario public health units (PHUs). Sex-specific and, when available, age-standardized regional rates of smoking, obesity, and racial/ethnic diversity were calculated at the public health unit level using national census and survey data available from Public Health Ontario.^4,31–33^ Racial/ethnic diversity was defined as the estimated regional proportion of individuals who self-identified as Black, South Asian, Chinese, Filipino, Latin American, Arab, Southeast Asian, West Asian, Korean, and Japanese according to national Census data.^34^ The receipt of influenza vaccination within the past year was determined from the OHIP and ODB databases among eligible individuals.

### Outcomes

Our outcome of interest was SARS-CoV-2 infection, determined by reverse transcription polymerase chain reaction (RT-PCR) testing. As individuals could undergo multiple tests over the study period, if any test was positive, they were classified as having the outcome. Otherwise, individuals testing negative for SARS-CoV-2 in all tests comprised the uninfected group.

### Statistical Analysis

Sociodemographic and clinical characteristics between individuals with and without SARS-CoV-2 infection were compared using chi-square tests for categorical variables and one-way analysis of variance for continuous variables. These comparisons were stratified by LTC residency status.

To evaluate the association between baseline characteristics and risk of SARS-CoV-2 infection, we fit multivariable logistic regression models to determine the odds ratio (OR) of testing positive. Since baseline health status and congregate living arrangements of LTC residents differ substantially from community-dwelling individuals, multivariable models were constructed separately for each group of patients. We regressed the outcome (testing positive vs. testing negative) on sociodemographic and clinical characteristics, incorporating PHU-specific random effects to account for clustering of individuals within communities. In these multivariable analyses, we adjusted for calendar week of testing, sociodemographic factors (age, sex, rural/urban residence, immigrant status, neighborhood income quintile, and regional racial/ethnicity diversity rate), and the aforementioned clinical risk factors. An interaction term was introduced to examine for heterogeneity in the association of age with the likelihood of SARS-CoV-2 infection over testing week. Given significant heterogeneity was detected among community-dwelling individuals, we stratified reporting of results into two time periods, one prior to the peak of the first wave of the pandemic and one following this period.

Sociodemographic and clinical risk factors that were independently associated with SARS-CoV-2 infection were then used to calculate an integer risk score for each individual by summing the number of risk factors the individual had. Separately for the periods prior to and following the peak of the first wave of the pandemic, we then calculated absolute infection rates and the odds ratio of SARS-CoV-2 infection in Ontario stratified by quartiles of regional rates of racial/ethnicity diversity (Ontario median 4.0%, interquartile range, 2.5-17.6%; Supplemental Figure 3). Associations of the integer scores with SARS-CoV-2 infection were assessed using logistic regression models. As few individuals had no risk factors following the peak of the pandemic, we combined the presence of 0 to 1 risk factors as the lowest risk group for the post-peak period. Trend in infection risk with increasing integer score was assessed using the Cochrane-Armitage trend test. All p-values were 2-sided with p<0.05 considered significant. The use of data in this project was authorized under section 45 of Ontario’s Personal Health Information Protection Act, which does not require review by a Research Ethics Board for use of anonymized data. The study followed the Strengthening the Reporting of Observational Studies in Epidemiology (STROBE) reporting guideline. All data were analyzed at ICES using SAS version 9.4 (SAS Institute, Cary, NC).

## Results

A description of the number of study participants and reasons for inclusion and exclusion are summarized in Supplemental Figure 2. Overall, among 644,102 eligible patients tested between January 1 and June 12, 2020, 25,446 (4.0%) adults tested positive for SARS-COV-2 infection. Among 578,263 community-dwelling individuals, 20,524 (3.5%) people tested positive, while among 65,839 LTC residents, 4,922 (7.5%) tested positive. The weekly number and rate of community-dwelling individuals first testing positive for SARS-COV-2 by age group is presented in Figure 1. The weekly number of community-dwelling individuals tested for SARS-CoV-2, and share of tests that were positive test, stratified by age group is presented in Supplemental Figure 4. During the first wave, the peak number and proportion of individuals with SARS-CoV-2 infection occurred during the week starting April 12, 2020, 3 weeks following a province-wide lockdown (Supplemental Figure 1).^35^

**Figure 1.**
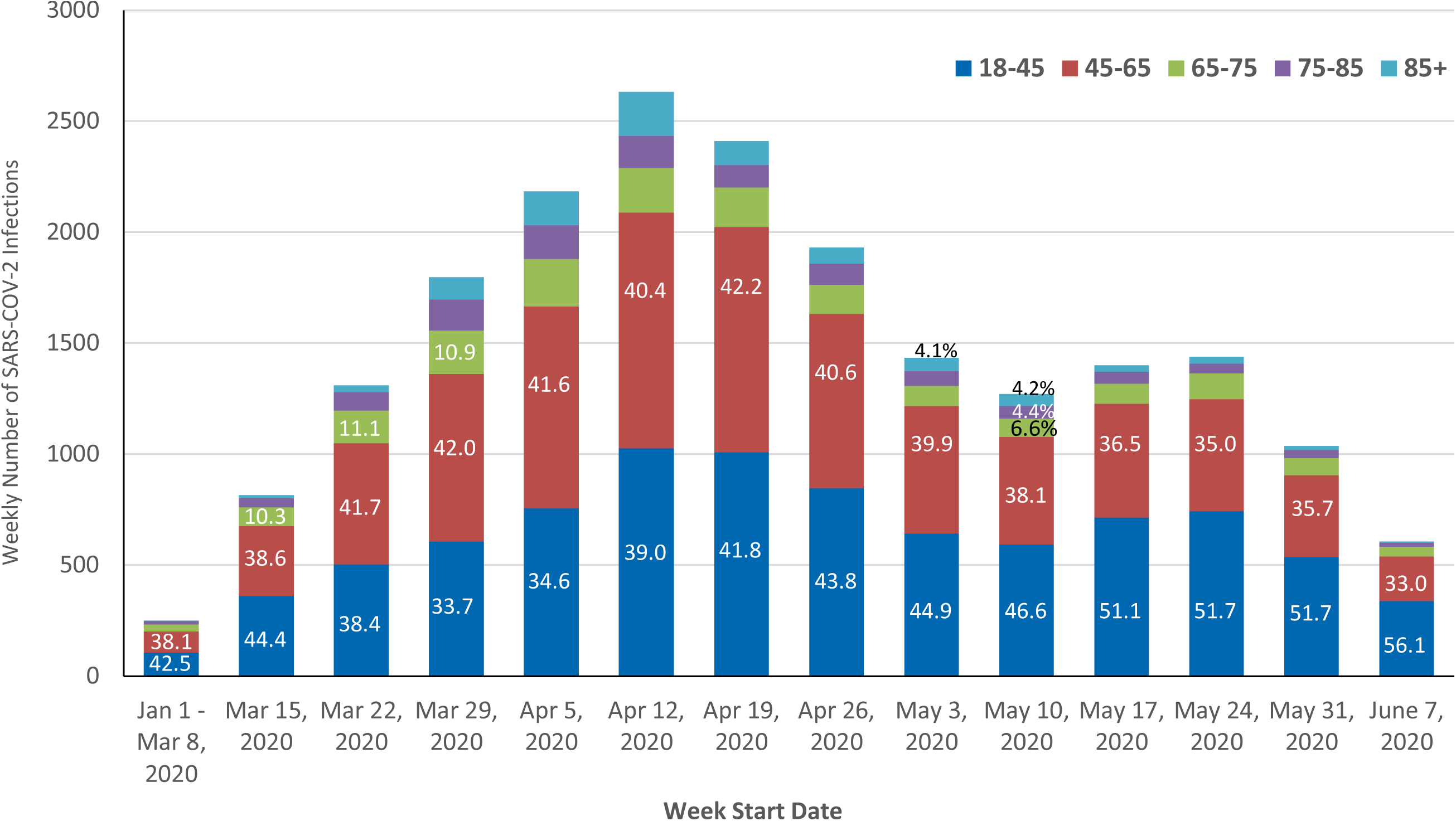
Weekly number of community-dwelling individuals with SARS-CoV-2 infection stratified by age group during the first wave of the pandemic in Ontario, Canada. Time represented by the start of the calendar week with the weeks of January 1 through March 8 consolidated given low initial infection counts. Age groups: 18-45 years (blue); 45-65 years (orange); 65-75 years (green); 75-85 years (yellow); 85 years and older (grey). The proportion of weekly counts attributable to an age group is represented in each column. The peak of the first wave of the pandemic in regard to community-dwelling infections in Ontario correlated with the week of April 12, 2020.

Baseline sociodemographic and clinical characteristics among community-dwelling individuals with and without SARS-CoV-2 infection are presented in Table 1. Compared with individuals without SARS-CoV-2 infection, community-dwelling individuals with SARS-CoV-2 infection were overall younger, more frequently male, immigrants, and residing in racially/ethnically diverse, large, urban, low-income communities. Individuals with SARS-CoV-2 infection had higher rates of diabetes, but otherwise had lower rates of most clinical comorbidities and lower rates of recent hospitalization or ED visits. Characteristics among LTC residents with SARS-CoV-2 infection are presented in Supplemental Table 2. Among individuals residing in LTC, those infected with SARS-CoV-2 were more frequently frail and had higher rates of recent hospitalization or ED visits, cardiovascular risk factors and comorbidities, and lung disease. Sociodemographic trends were similar to the community-dwelling population.

**Table 1.** Baseline Characteristics of Community-Dwelling Patients with and without SARS-CoV-2 Infection During the First Wave of the Pandemic in Ontario, Canada.

| Characteristic |  | Negative Test<br>(n=557,739) | Positive Test<br>(n=20,524) | SD | P-value |
| --- | --- | --- | --- | --- | --- |
| Week of testing (start date) | 0-10 (Jan. 1 – Mar. 8) | 8,477 (1.5%) | 252 (1.2%) | 0.03 | <0.001 |
|  | 11 (Mar. 15) | 16,069 (2.9%) | 816 (4.0%) | 0.06 |  |
|  | 12 (Mar. 22) | 15,203 (2.7%) | 1,311 (6.4%) | 0.18 |  |
|  | 13 (Mar. 29) | 15,921 (2.9%) | 1,798 (8.8%) | 0.25 |  |
|  | 14 (Apr. 5) | 19,749 (3.5%) | 2,184 (10.6%) | 0.28 |  |
|  | 15 (Apr. 12) | 34,058 (6.1%) | 2,632 (12.8%) | 0.23 |  |
|  | 16 (April 19) | 42,864 (7.7%) | 2,410 (11.7%) | 0.14 |  |
|  | 17 (April 26) | 52,234 (9.4%) | 1,932 (9.4%) | 0 |  |
|  | 18 (May 3) | 55,546 (10.0%) | 1,435 (7.0%) | 0.11 |  |
|  | 19 (May 10) | 45,758 (8.2%) | 1,272 (6.2%) | 0.08 |  |
|  | 20 (May 17) | 33,468 (6.0%) | 1,400 (6.8%) | 0.03 |  |
|  | 21 (May 24) | 71,835 (12.9%) | 1,439 (7.0%) | 0.20 |  |
|  | 22 (May 31) | 78,056 (14.0%) | 1,037 (5.1%) | 0.31 |  |
|  | 23 (June 7) | 68,501 (12.3%) | 606 (3.0%) | 0.36 |  |
| Age, years |  | 51 (35-64) | 49 (34-61) | 0.09 | <0.001 |
| Age group, years | 18-45 | 230,307 (41.3%) | 8,789 (42.8%) | 0.03 | <0.001 |
|  | 45-65 | 195,925 (35.1%) | 8,130 (39.6%) | 0.09 |  |
|  | 65-75 | 59,943 (10.7%) | 1,683 (8.2%) | 0.09 |  |
|  | 75-85 | 40,276 (7.2%) | 1,053 (5.1%) | 0.09 |  |
|  | ≥85 | 31,288 (5.6%) | 869 (4.2%) | 0.06 |  |
| Sex | Female | 344,040 (61.7%) | 11,186 (54.5%) | 0.15 | <0.001 |
|  | Male | 213,699 (38.3%) | 9,338 (45.5%) | 0.15 |  |
| Income quintile | 1 | 122,004 (21.9%) | 5,827 (28.4%) | 0.15 | <0.001 |
|  | 2 | 115,955 (20.8%) | 4,343 (21.2%) | 0.01 |  |
|  | 3 | 111,368 (20.0%) | 4,168 (20.3%) | 0.01 |  |
|  | 4 | 106,160 (19.0%) | 3,361 (16.4%) | 0.07 |  |
|  | 5 | 102,252 (18.3%) | 2,825 (13.8%) | 0.12 |  |
| Community size | <10,000 | 63,441 (11.4%) | 743 (3.6%) | 0.30 | <0.001 |
|  | 10,000-100,000 | 53,810 (9.6%) | 832 (4.1%) | 0.22 |  |
|  | 100,000-500,000 | 134,283 (24.1%) | 2,543 (12.4%) | 0.31 |  |
|  | 500,000-1.5 million | 87,383 (15.7%) | 2,427 (11.8%) | 0.11 |  |
|  | ≥1.5 million | 218,822 (39.2%) | 13,979 (68.1%) | 0.61 |  |
| Canadian immigrant |  | 94,187 (16.9%) | 7,836 (38.2%) | 0.49 | <0.001 |
| Regional smoking rate, % |  | 12 (9-17) | 12 (9-13) | 0.41 | <0.001 |
| Regional obesity rate, % |  | 21 (15-24) | 16 (15-22) | 0.47 | <0.001 |
| Regional racial diversity rate, % |  | 19 (7-51) | 51 (25-52) | 0.65 | <0.001 |

Table 1 Continued. Baseline Characteristics of Community-Dwelling Patients with and without SARS-CoV-2 Infection During the First Wave of the Pandemic in Ontario, Canada.
| Characteristic |  | Negative Test<br>(n=557,739) | Positive Test<br>(n=20,524) | SD | P-value |
| --- | --- | --- | --- | --- | --- |
| **Clinical risk factors** |  |  |  |  |  |
| Hypertension |  | 174,916 (31.4%) | 6,317 (30.8%) | 0.01 | 0.077 |
| Hypertension duration,<br>years |  | 13 (7-20) | 12 (6-19) | 0.14 | <.001 |
| Diabetes |  | 66,994 (12.0%) | 2,916 (14.2%) | 0.07 | <.001 |
| Coronary artery disease |  | 24,352 (4.4%) | 619 (3.0%) | 0.07 | <.001 |
| Heart failure |  | 11,921 (2.1%) | 308 (1.5%) | 0.05 | <.001 |
| Stroke |  | 4,576 (0.8%) | 158 (0.8%) | 0.01 | 0.429 |
| Atrial fibrillation |  | 24,555 (4.4%) | 631 (3.1%) | 0.07 | <.001 |
| Chronic kidney disease |  | 27,673 (5.0%) | 854 (4.2%) | 0.04 | <.001 |
| HIV |  | 1,630 (0.3%) | 56 (0.3%) | 0 | 0.613 |
| Organ transplantation |  | 1,390 (0.2%) | 31 (0.2%) | 0.02 | 0.005 |
| Cancer |  | 19,324 (3.5%) | 401 (2.0%) | 0.09 | <.001 |
| Liver disease |  | 3,502 (0.6%) | 78 (0.4%) | 0.04 | <.001 |
| Lung disease |  | 129,702 (23.3%) | 3,742 (18.2%) | 0.12 | <.001 |
| Hospitalization or ED visits<br>in 2019 | 0 | 261,024 (46.8%) | 11,254 (54.8%) | 0.16 | <.001 |
|  | 1-2 | 186,021 (33.4%) | 6,522 (31.8%) | 0.03 |  |
|  | 3+ | 110,694 (19.8%) | 2,748 (13.4%) | 0.17 |  |
| Frailty |  | 32,041 (5.7%) | 995 (4.8%) | 0.04 | <.001 |
| Influenza vaccination in<br>2019 |  | 163,565 (29.3%) | 4,792 (23.3%) | 0.14 | <.001 |

Continuous variables are reported as median (interquartile range). Abbreviations: ED: emergency department; HIV: human immunodeficiency virus; SD: standardized difference.

Independent predictors of SARS-CoV-2 infection among community-dwelling individuals are presented in Table 2. Results are stratified into the period prior to (Table 2a) and following (Table 2b) the peak of the first wave of the pandemic in the province given the detection of significant heterogeneity in the risk of SARS-CoV-2 infection by age across time (P-interaction <0.0001). During the period leading up to the peak of the first wave of the pandemic (Table 2a), the likelihood of SARS-CoV-2 infection progressively increased across age groups (age 45-65, OR 1.30, 95% CI 1.24-1.37; age 65-75, OR 1.38, 95% CI 1.26-1.51; age 75-85, OR 1.46, 95% CI 1.30-1.63; age 85 and older, OR 1.60, 95% CI 1.41-1.81) compared with the youngest individuals aged 18-45 years. Other independent risk factors for SARS-CoV-2 infection included male sex (OR 1.45, 95% CI 1.31-1.60), residing in the lowest quintile of neighborhood income (OR 1.09, 95% CI 1.01-1.17), residing in more racially/ethnically diverse communities (OR per 1% increase in regional racial/ethnic diversity 1.02, 95% CI 1.01-1.03), immigration to Canada (OR 1.53, 95% CI 1.45-1.61), frailty (OR 1.31, 95% CI 1.01-1.25), hypertension (OR 1.10, 95% CI 1.04-1.17), and diabetes (OR 1.12, 95% CI 1.05-1.20).

**Table 2.** Predictors of SARS-CoV-2 infection prior to (a) and following (b) the peak of the first wave of the pandemic among community-dwelling individuals in Ontario, Canada.

**a) Weeks of January 1 - April 12, 2020**
| Characteristic |  | OR (95% CI) | P-value |
| --- | --- | --- | --- |
| Week of testing (start date) | 0-10 (Jan. 1 – Mar. 8) | Referent | <0.0001 |
|  | 11 (March 15) | 1.95 (1.69-2.26) |  |
|  | 12 (March 22) | 3.63 (3.16-4.18) |  |
|  | 13 (March 29) | 4.87 (4.25-5.58) |  |
|  | 14 (April 5) | 4.69 (4.10-5.37) |  |
|  | 15 (April 12) | 2.99 (2.62-3.42) |  |
| Age group, years | 18-45 | Referent | <0.0001 |
|  | 45-65 | 1.30 (1.24-1.37) |  |
|  | 65-75 | 1.38 (1.26-1.51) |  |
|  | 75-85 | 1.46 (1.30-1.63) |  |
|  | ≥85 | 1.60 (1.41-1.81) |  |
| Sex | Female | Referent |  |
|  | Male | 1.45 (1.31-1.60) | <0.0001 |
| Rural dwelling |  | 0.96 (0.86-1.09) | 0.56 |
| Income quintile | 1 | 1.09 (1.01-1.17) |  |
|  | 2 | 0.96 (0.89-1.03) |  |
|  | 3 | 1.05 (0.98-1.13) |  |
|  | 4 | 1.01 (0.94-1.08) |  |
|  | 5 | Referent | 0.005 |
| Canadian immigrant |  | 1.53 (1.45-1.61) | <0.0001 |
| Frailty |  | 1.13 (1.01-1.25) | 0.03 |
| Hospitalization or ED visits in 2019 | 0 | Referent | <0.0001 |
|  | 1-2 | 0.77 (0.73-0.81) |  |
|  | 3+ | 0.54 (0.51-0.58) |  |
| Hypertension |  | 1.10 (1.04-1.17) | 0.001 |
| Diabetes |  | 1.12 (1.05-1.20) | 0.001 |
| Coronary artery disease |  | 0.78 (0.69-0.88) | <0.0001 |
| Heart failure |  | 0.96 (0.81-1.14) | 0.64 |
| Stroke |  | 1.13 (0.89-1.42) | 0.31 |
| Atrial fibrillation |  | 0.87 (0.77-0.99) | 0.03 |
| Chronic kidney disease |  | 0.83 (0.74-0.93) | 0.001 |
| HIV |  | 0.64 (0.41-0.98) | 0.04 |
| Organ transplantation |  | 0.61 (0.36-1.01) | 0.05 |
| Cancer |  | 0.70 (0.61-0.81) | <0.0001 |
| Liver disease |  | 0.66 (0.47-0.91) | 0.01 |
| Lung disease |  | 0.83 (0.78-0.88) | <0.0001 |
| Influenza vaccination in 2019 |  | 0.98 (0.93-1.03) | 0.41 |
| Regional smoking rate, per 1% |  | 1.00 (0.98-1.02) | 0.65 |
| Regional obesity rate, per 1% |  | 0.98 (0.97-0.995) | 0.003 |
| Regional racial diversity rate, per 1% |  | 1.02 (1.01-1.03) | 0.002 |

b) Weeks of April 19 – June 7, 2020
| Characteristic |  | OR (95% CI) | P-value |
| --- | --- | --- | --- |
| Week of testing (start date) | 16 (April 19) | Referent | <0.0001 |
|  | 17 (April 26) | 0.65 (0.62-0.70) |  |
|  | 18 (May 3) | 0.49 (0.46-0.52) |  |
|  | 19 (May 10) | 0.49 (0.46-0.53) |  |
|  | 20 (May 17) | 0.68 (0.64-0.73) |  |
|  | 21 (May 24) | 0.34 (0.32-0.36) |  |
|  | 22 (May 31) | 0.22 (0.20-0.24) |  |
|  | 23 (June 7) | 0.15 (0.14-0.17) |  |
| Age group (years) | 18-45 | 1.58 (1.40-1.80) | <0.0001 |
|  | 45-65 | 1.43 (1.27-1.62) |  |
|  | 65-75 | 1.23 (1.08-1.40) |  |
|  | 75-85 | 1.11 (0.97-1.28) |  |
|  | ≥85 | Referent |  |
| Sex | Female | Referent |  |
|  | Male | 1.67 (1.52-1.83) | <0.0001 |
| Rural dwelling |  | 0.90 (0.79-1.04) | 0.14 |
| Income quintile | 1 | 2.01 (1.87-2.16) |  |
|  | 2 | 1.53 (1.43-1.65) |  |
|  | 3 | 1.46 (1.36-1.57) |  |
|  | 4 | 1.27 (1.17-1.37) |  |
|  | 5 | Referent | <0.0001 |
| Canadian immigrant |  | 1.82 (1.75-1.90) | <0.0001 |
| Frailty |  | 0.94 (0.84-1.06) | 0.32 |
| Hospitalization or ED visits in 2019 | 0 | Referent | <0.0001 |
|  | 1-2 | 0.97 (0.93-1.01) |  |
|  | 3+ | 0.82 (0.77-0.87) |  |
| Hypertension |  | 1.07 (1.01-1.12) | 0.02 |
| Diabetes |  | 1.32 (1.24-1.40) | <0.0001 |
| Coronary artery disease |  | 0.82 (0.73-0.93) | 0.002 |
| Heart failure |  | 0.86 (0.71-1.05) | 0.14 |
| Stroke |  | 1.05 (0.82-1.34) | 0.71 |
| Atrial fibrillation |  | 0.92 (0.80-1.04) | 0.17 |
| Chronic kidney disease |  | 0.70 (0.62-0.78) | <0.0001 |
| HIV |  | 0.67 (0.48-0.95) | 0.03 |
| Organ transplantation |  | 0.71 (0.42-1.20) | 0.20 |
| Cancer |  | 0.53 (0.45-0.62) | <0.0001 |
| Liver disease |  | 0.65 (0.47-0.90) | 0.009 |
| Lung disease |  | 0.90 (0.85-0.95) | <0.0001 |
| Influenza vaccination in 2019 |  | 0.78 (0.74-0.82) | <0.0001 |
| Regional smoking rate, per 1% |  | 0.99 (0.97-1.01) | 0.25 |
| Regional obesity rate, per 1% |  | 1.01 (1.00-1.02) | 0.30 |
| Regional racial diversity rate, per 1% |  | 1.05 (1.03-1.06) | <0.0001 |

Odds ratios were determined from a multivariable logistic regression model that incorporated PHU-specific random effects. For continuous variables, results are per 1-unit increase in rate. There was significant heterogeneity in the overall model between age and week of testing (P-interaction <0.0001), hence results are presented stratified prior to and following the peak of the first wave of the pandemic. Abbreviations: CI: confidence intervals; ED: emergency department; HIV: human immunodeficiency virus; OR; odds ratio.

Following the peak of the first wave of the pandemic the likelihood of SARS-CoV-2 infection across age groups in community-dwelling individuals reversed (Supplemental Figure 4). Thereafter, the oldest individuals aged ≥ 85 years had the lowest likelihood SARS-CoV-2 infection (Table 2b), with a progressive increase in infection risk as age declined compared with individuals aged ≥ 85 years (age 75-85, OR 1.11, 95% CI 0.97-1.28; age 65-75, OR 1.23, 95% CI 1.08-1.40; age 45-65, OR 1.43, 95% CI 1.27-1.62; and age 18-45, OR 1.58, 95% CI 1.40-1.80). Additionally, there was a progressive increased risk of SARS-CoV-2 infection across all lower quintiles of neighborhood income compared with the highest quintile (quintile 1, OR 2.01, 95% CI 1.87-2.16; quintile 2, OR 1.53, 95% CI 1.43-1.65; quintile 3, OR 1.46, 95% CI 1.36-1.57; quintile 4, OR 1.27, 95% CI 1.17-1.37). Other independent risk factors included male sex (OR 1.67, 95% CI 1.52-1.83), residing in more racially/ethnically diverse communities (OR per 1% increase 1.05, 95% CI 1.03-1.06), immigration to Canada (OR 1.82, 95% CI 1.75-1.90), history of hypertension (OR 1.07, 95% CI 1.01-1.12), and history of diabetes (OR 1.32, 95% CI 1.24-1.40).

In contrast to predictors in the general community, there was no heterogeneity over time in the adjusted risk associated with age and SARS-CoV-2 infection among LTC residents (Supplemental Table 3). Moreover, in this population, age and sex were not independent predictors for SARS-CoV-2 infection whereas residing in non-rural, more racially diverse communities remained significantly associated with testing positive.

The absolute and relative risk of SARS-CoV-2 infection among community-dwelling individuals according to the number of independent risk factors identified above (e.g., age category, male sex, residing in a lower income neighborhood, Canadian immigrant status, hypertension, diabetes, and, prior to the peak of the pandemic, a history of frailty), degree of regional racial/ethnic diversity, and time period are shown in Figure 2 and Table 3. Prior to the peak of the pandemic, SARS-CoV-2 infection rates were generally greater across communities with more racial/ethnic diversity and among individuals with a higher number of risk factors such that individuals living in the most racially/ethnically diverse communities without any other risk factors had a similar rate of infection as individuals living in the least racially/ethnically diverse communities with 3 or more risk factors (Figure 2a). Individuals with 1, 2, or ≥3 risk factors had progressively higher odds of SARS-CoV-2 infection compared with individuals without risk factors across regions of racial/ethnic diversity.

**Table 3.** Rates and odds ratios of SARS-CoV-2 infection by number of risk factors and degree of regional racial/ethnic diversity, prior to and following the peak of the first wave of the pandemic among community-dwelling individuals in Ontario, Canada.

**Prior to the Peak of the First Wave (Weeks of January 1 - April 12, 2020)**
| Racial/Ethnic Diversity Quartile 1<br>($\leq 2.5\%$; least diverse) | COVID-19 Positive<br>(n=390) (4.8%) | Total<br>(n=8,123) | OR (95% CI) | P-trend |
| --- | --- | --- | --- | --- |
| Number of risk factors |  |  |  |  |
| 0 | 64 (4.2%) | 1,518 | Referent | 0.017 |
| 1 | 105 (4.3%) | 2,451 | 1.02 (0.74-1.40) |  |
| 2 | 98 (4.9%) | 1,989 | 1.18 (0.85-1.63) |  |
| 3+ | 123 (5.7%) | 2,165 | 1.37 (1.004-1.87) |  |
| Racial/Ethnic Diversity Quartile 2<br>(2.6-4.0%) | COVID-19 Positive<br>(n=348) (3.1%) | Total<br>(n=11,333) |  |  |
| Number of risk factors |  |  |  |  |
| 0 | 49 (2.2%) | 2,263 | Referent | 0.308 |
| 1 | 118 (3.4%) | 3,445 | 1.60 (1.14-2.25) |  |
| 2 | 99 (3.7%) | 2,676 | 1.74 (1.23-2.46) |  |
| 3+ | 82 (2.8%) | 2,949 | 1.29 (0.90-1.85) |  |
| Racial/Ethnic Diversity Quartile 3<br>(4.1-17.6%) | COVID-19 Positive<br>(n=1,184) (5.5%) | Total<br>(n=21,560) |  |  |
| Number of risk factors |  |  |  |  |
| 0 | 142 (3.1%) | 4,556 | Referent | <0.001 |
| 1 | 336 (4.8%) | 6,960 | 1.58 (1.29-1.93) |  |
| 2 | 307 (5.8%) | 5,337 | 1.90 (1.55-2.32) |  |
| 3+ | 399 (6.8%) | 5,891 | 2.26 (1.86-2.75) |  |
| Racial/Ethnic Diversity Quartile 4<br>($\geq 17.7\%$; most diverse) | COVID-19 Positive<br>(n=7,071) (9.3%) | Total<br>(n=76,270) | | |
| Number of risk factors |  |  |  |  |
| 0 | 704 (5.8%) | 12,052 | Referent | <0.001 |
| 1 | 1,684 (7.9%) | 21,390 | 1.38 (1.26-1.51) |  |
| 2 | 1,897 (9.8%) | 19,320 | 1.76 (1.61-1.92) |  |
| 3+ | 2,786 (11.9%) | 23,508 | 2.17 (1.99-2.36) |  |

**Following the First Wave Peak (Weeks of April 19 - June 7, 2020)**
| Racial/Ethnic Diversity Quartile 1<br>(≤2.5%; least diverse) | COVID-19 Positive<br>(n=165) (0.5%) | Total<br>(n=36,173) | OR (95% CI) | P-trend |
| --- | --- | --- | --- | --- |
| Number of risk factors |  |  |  |  |
| 0-1 | 15 (0.5%) | 2,785 | Referent | 0.32 |
| 2 | 61 (0.4%) | 15,899 | 0.71 (0.40-1.25) |  |
| 3 | 59 (0.5%) | 11,772 | 0.93 (0.53-1.64) |  |
| 4+ | 30 (0.5%) | 5,717 | 0.97 (0.52-1.81) |  |
| Racial/Ethnic Diversity Quartile 2<br>(2.6-4.0%) | COVID-19 Positive<br>(n=204) (0.3%) | Total<br>(n=59,811) |  |  |
| Number of risk factors |  |  |  |  |
| 0-1 | 15 (0.3%) | 4,509 | Referent | 0.63 |
| 2 | 79 (0.3%) | 25,811 | 0.92 (0.53-1.60) |  |
| 3 | 80 (0.4%) | 19,524 | 1.23 (0.71-2.14) |  |
| 4+ | 30 (0.3%) | 9,967 | 0.91 (0.49-1.68) |  |
| Racial/Ethnic Diversity Quartile 3<br>(4.1-17.6%) | COVID-19 Positive<br>(n=924) (1.0%) | Total<br>(n=91,689) |  |  |
| Number of risk factors |  |  |  |  |
| 0-1 | 73 (0.9%) | 7,813 | Referent | <0.001 |
| 2 | 330 (0.9%) | 37,713 | 0.94 (0.73-1.21) |  |
| 3 | 334 (1.1%) | 30,886 | 1.16 (0.90-1.50) |  |
| 4+ | 187 (1.2%) | 15,277 | 1.31 (1.001-1.73) |  |
| Racial/Ethnic Diversity Quartile 4<br>(≥17.7%; most diverse) | COVID-19 Positive<br>(n=10,238) (3.8%) | Total<br>(n=272,120) |  |  |
| Number of risk factors |  |  |  |  |
| 0-1 | 258 (1.3%) | 19,583 | Referent | <0.001 |
| 2 | 2,128 (2.5%) | 86,572 | 1.89 (1.66-2.15) |  |
| 3 | 3,928 (3.9%) | 99,931 | 3.07 (2.70-3.48) |  |
| 4+ | 3,924 (5.9%) | 66,034 | 4.73 (4.17-5.37) |  |

Number of potential risk factors following the first wave were comprised of male sex, age < 85 years, neighborhood income quintiles 1-4, Canadian immigrant, hypertension, and diabetes. Region was categorized by quartiles of community racial/ethnic diversity rate.

**Figure 2.**
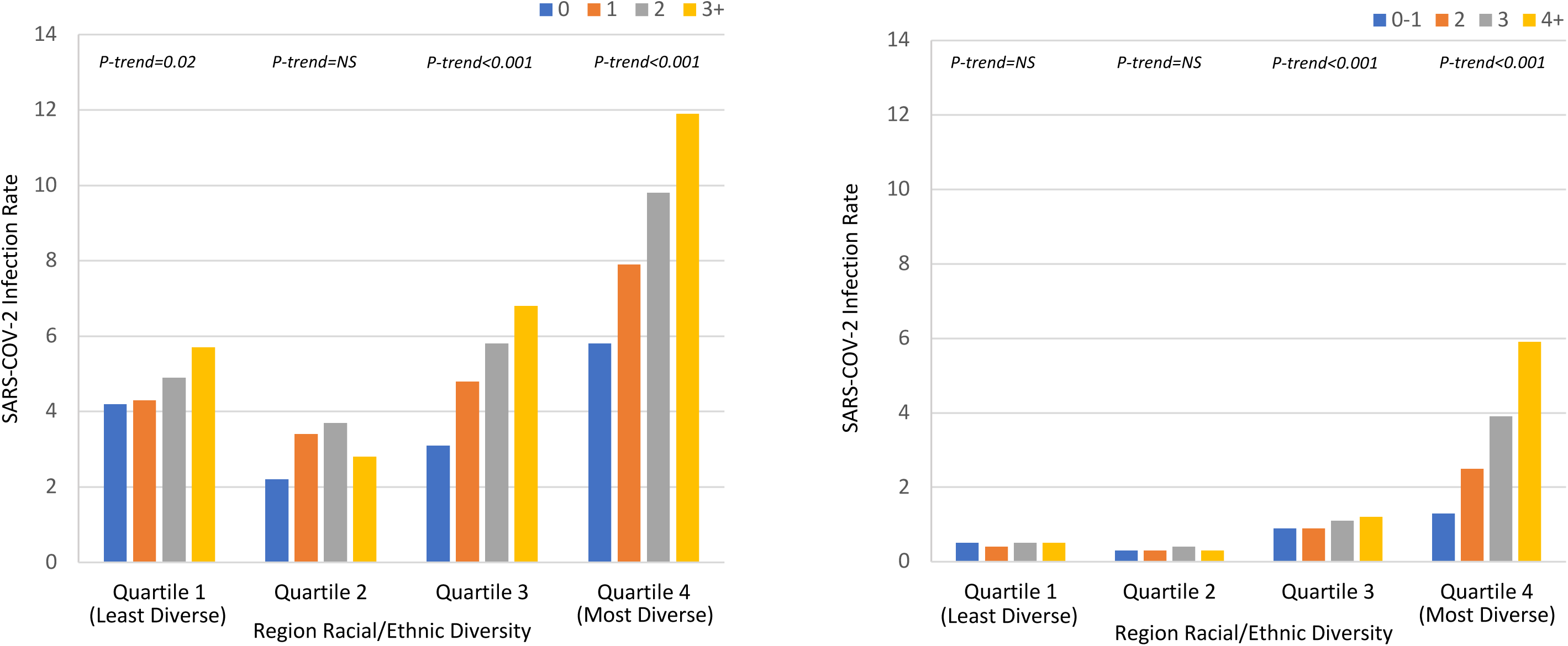
SARS-CoV-2 infection rates by number of risk factors and degree of regional racial/ethnic diversity, prior to and following the peak of the first wave of the pandemic in Ontario, Canada. a) Weeks of January 1 – April 12, 2020; b) Weeks of April 19 – June 7, 2020. Individuals were classified according to the number of potential risk factors present. Risk factors prior to the first wave were comprised of: male sex, age > 45 years, lowest quintile of neighborhood income, Canadian immigrant, history of frailty, hypertension, and diabetes; following the first wave: male sex, age < 85 years, neighborhood income quintiles 1-4, Canadian immigrant, hypertension, and diabetes. Region was categorized by quartiles of community racial/ethnic diversity rate.

Following the peak of the first wave of the pandemic in mid-April, there was a reduction in infection rates across Ontario, however there was less impact in regions with higher degrees of racial/ethnic diversity. Additionally, while accumulation of more risk factors remained associated with a higher risk of SARS-CoV-2 infection in the most racially/ethnically diverse communities, the risk factors now included all age groups < 85 years (Figure 2b; Table 3).

Individuals living in the most racially/ethnically diverse communities with 2, 3, or ≥4 risk factors had ORs of 1.89, 3.07, and 4.73 for SARS-CoV-2 infection compared to lower risk individuals in their community with 0-1 risk factors. In contrast, in the least racially/ethnically diverse communities, there was little to no gradient in infection rates across risk strata.

## Discussion

We observed three dynamic factors during the first wave of the pandemic associated with SARS-CoV-2 infection among community-dwelling individuals in Ontario that merit consideration for risk determination, response planning, and health policy. First, we detected significant time-varying heterogeneity in the association between age and the odds of SARS-CoV-2 infection. Initially, an incremental increase in age was independently associated with a higher odds of SARS-CoV-2 infection, a biologic risk factor representing a more vulnerable population susceptible to viral infection. After the implementation of public health measures in late March 2020, the association reversed with the odds of SARS-CoV-2 infection increasing across progressively younger age groups compared with community-dwelling individuals 85 years and older. Second, the number of independent risk factors was associated with a stepwise increase in the odds of SARS-CoV-2 infection. Across Ontario, there was an initial increased odds of infection associated with the presence of more clinical risk factors (such as diabetes and hypertension), which was accentuated in regions with higher racial/ethnic diversity. After public health measures were implemented, the absolute and relative risk associated with the accumulation of risk factors diminished overall. However, among the regions of Ontario with the highest rates of racial/ethnic diversity, the relative odds of infection associated with a higher number of risk factors remained present. Third, we observed that the risk of SARS-CoV-2 infection was associated with higher regional racial/ethnic diversity. Following public health measures, regional racial/ethnic diversity remained independently associated with higher odds of SARS-CoV-2 infection, though absolute rates were reduced.

We analyzed cumulative risk stratified across quartiles of community racial/ethnic diversity to ascertain to what degree the presence of an increasing burden of sociodemographic and clinical risk factors was predictive of SARS-CoV-2 infection independent of residential factors. Regions of Ontario with the highest racial/ethnic diversity correlate with the largest sized communities and the highest neighborhood density, household crowding, and deprivation.^36^ Lower infection rates in the period following the pandemic’s initial peak correlated with implementation of national and provincial restrictions, including restrictions in international travel and closure of schools and non-essential businesses. These data suggest that these broad public-health interventions are associated with changes in the likelihood of infection by age, but had little impact on the other risk factors driving virus transmission. These risk factors may represent characteristics of individuals at higher risk of exposure, those working in essential services or living in densely populated housing, or both, and not represent a higher biologic susceptibility to infection. Systemic and structural inequities in these determinants of health are likely associated with higher residential and occupational risk of viral transmission and reduced ability to comply with isolation orders. This data may help inform policy to protect more vulnerable populations including essential workers, such as the implementation of paid sick leave, targeted screening in heavily impacted neighborhoods, and “wrap-around” services, to ensure those infected have a safe place to isolate and not infect others in the home.

There are several strengths of our study. This is the first North American population-based analysis of the cumulative effect of clinical and sociodemographic risk factors stratified by community racial/ethnic diversity and time. This approach unmasked considerable differences in the age-related susceptibility to SARS-CoV-2 infection before and after the peak of the first wave of the pandemic, and the residual risk that remained despite broad public-health measures in large, urban, racially diverse regions of the province most impacted by COVID-19. These findings have implications for the effectiveness of current public health measures to restrict SARS-CoV-2 infection, particularly among young, male, immigrants living in lower socioeconomic neighborhoods who are likely working or living in conditions not conducive to adherence to these measures. There are important limitations to acknowledge as well. Access to testing for SARS-CoV-2 infection varied over time, including restriction of testing during the earlier periods to the highest risk patients; while some patients with severe COVID-19 illness may also have died before testing. Over time, testing capacity increased and broader testing occurred. As a result, our analysis was focused on individuals that underwent SARS-CoV-2 testing to reduce ascertainment bias. In addition, a small number of hospital-based SARS-CoV-2 test results were not available for this analysis, but this low percentage of missingness would not be expected to materially impact the results of available data. Since we did not have individual-level data on weight, smoking status, income, and race/ethnicity, we relied on community-level variables as proxies. During both time periods, a number of chronic conditions were significantly associated with a lower risk of SARS-CoV-2 infection, likely reflecting collider/screening bias among asymptomatic patients undergoing regular care.^37^

Despite the dramatic impact of a provincial lockdown, following the peak of the initial wave of the pandemic in early April, the highest likelihood of SARS-CoV-2 infection emerged among clusters of people represented by younger age, male sex, individuals that immigrated to Canada, with hypertension or diabetes residing in the most racially/ethnically diverse, urban, most socioeconomically disadvantaged communities of Ontario. Further efforts appear necessary to reduce the risk of SARS-CoV-2 infection among the highest risk individuals residing in these communities.

## Data Availability

The dataset from this study is held securely in coded form at ICES. While legal data sharing agreements between ICES and data providers (e.g., healthcare organizations and government) prohibit ICES from making the dataset publicly available, access may be granted to those who meet pre-specified criteria for confidential access, available at www.ices.on.ca/DAS.

## Acknowledgements

This study was supported by ICES, which is funded by an annual grant from the Ontario Ministry of Health and Long-Term Care (MOHLTC). Parts of this material are based on data and information compiled and provided by the MOHLTC, Cancer Care Ontario (CCO), Canadian Institute for Health Information (CIHI), and CorHealth Ontario. The analyses, conclusions, opinions and statements expressed herein are solely those of the authors and do not reflect those of the funding or data sources; no endorsement is intended or should be inferred. Parts of this material are based on data and/or information compiled and provided by Immigration, Refugees and Citizenship Canada (IRCC). However, the analyses, conclusions, opinions and statements expressed in the material are those of the author(s), and not necessarily those of IRCC. The Public Health Unit data used for this manuscript was adapted with the permission of Public Health Ontario. Public Health Ontario assumes no responsibility for the content of any publication resulting from translation/changes/adaptation of PHO documents by third parties. The authors acknowledge that the clinical registry data used in this publication is from participating hospitals through CorHealth Ontario, which serves as an advisory body to the MOHLTC, is funded by the MOHLTC, and is dedicated to improving the quality, efficiency, access and equity in the delivery of the continuum of adult cardiac, vascular and stroke services in Ontario, Canada. This study made use of the Johns Hopkins ACG® System (Version 10) to identify frailty conditions.

## Funding

This study received funding from a Canadian Institutes of Health Research (CIHR) Strategy for Patient-Oriented Research Innovative Clinical Trial Multi-year grant (MYG-151211), a Ted Rogers Centre for Heart Research Innovation Fund -COVID-19 Award, a Peter Munk Cardiac Care Innovation Fund, and a COVID-19 Rapid Research Funding Opportunity – Clinical Management and Health Systems Interventions from the Institute of Circulatory and Respiratory Health (ICRH)-CIHR operating grant: (VR4-172736). Dr. Udell is supported by a Government of Ontario Early Researcher Award (ER15-11-037), Women’s College Research Institute and Department of Medicine, Women’s College Hospital. Dr. Austin is supported by a Mid-Career Investigator Award from the Heart and Stroke Foundation. Dr. Goodman was supported by the Heart and Stroke Foundation of Ontario/University of Toronto Polo Chair. Dr. McAlister holds the Alberta Health Services Chair in Cardiovascular Outcomes Research. Dr. Lee is the Ted Rogers Chair in Heart Function Outcomes, University Health Network, University of Toronto.

## Disclosures

All authors have completed and submitted the ICMJE Form for Disclosure of Potential Conflicts of Interest.

## Supplemental Appendix

**Supplemental Figure 1.**
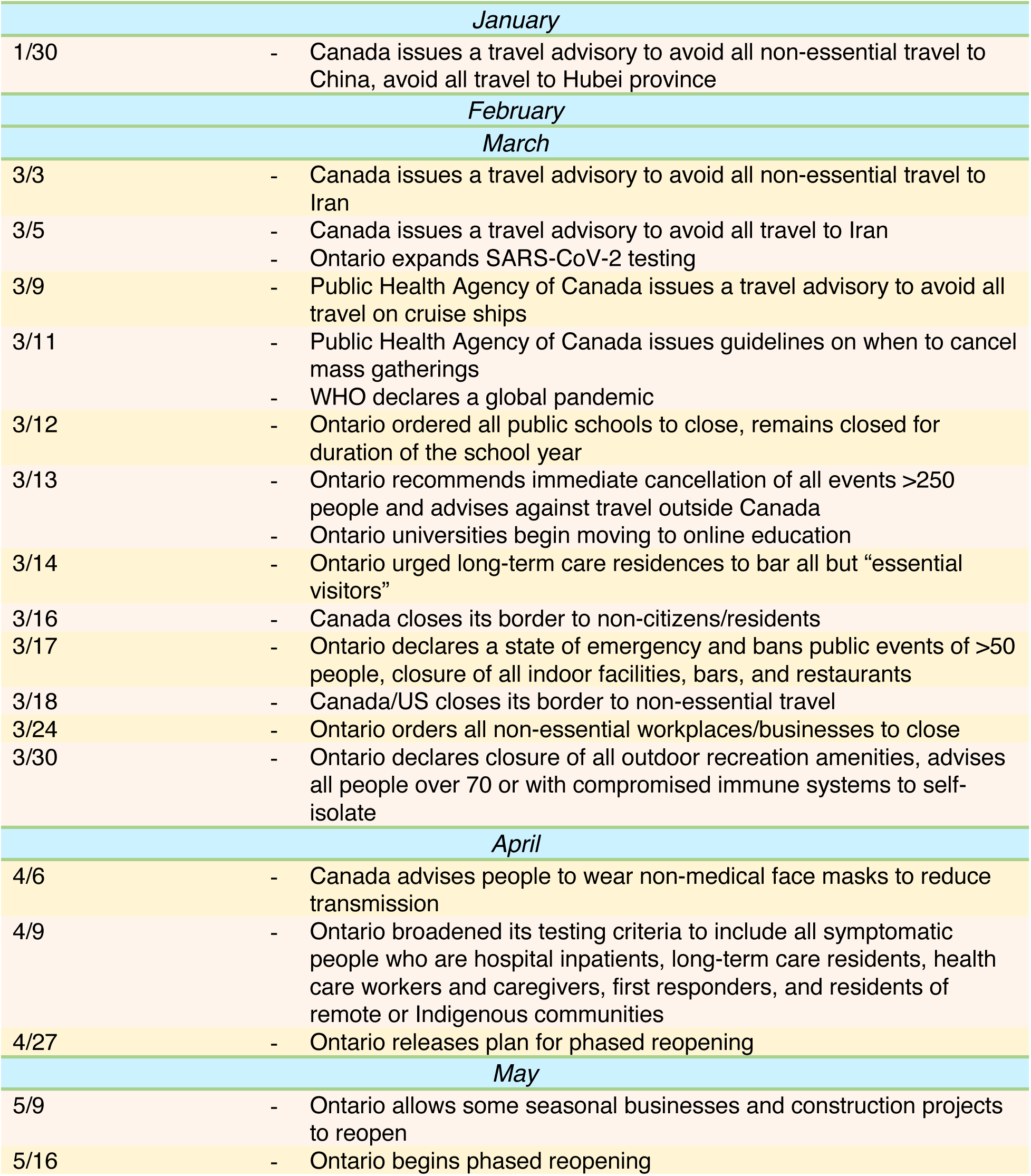
Timeline of Ontario/Canada’s First Wave Response to COVID-19 Until May 2020 (Vogel, 2020)

**Supplemental Figure 2.**
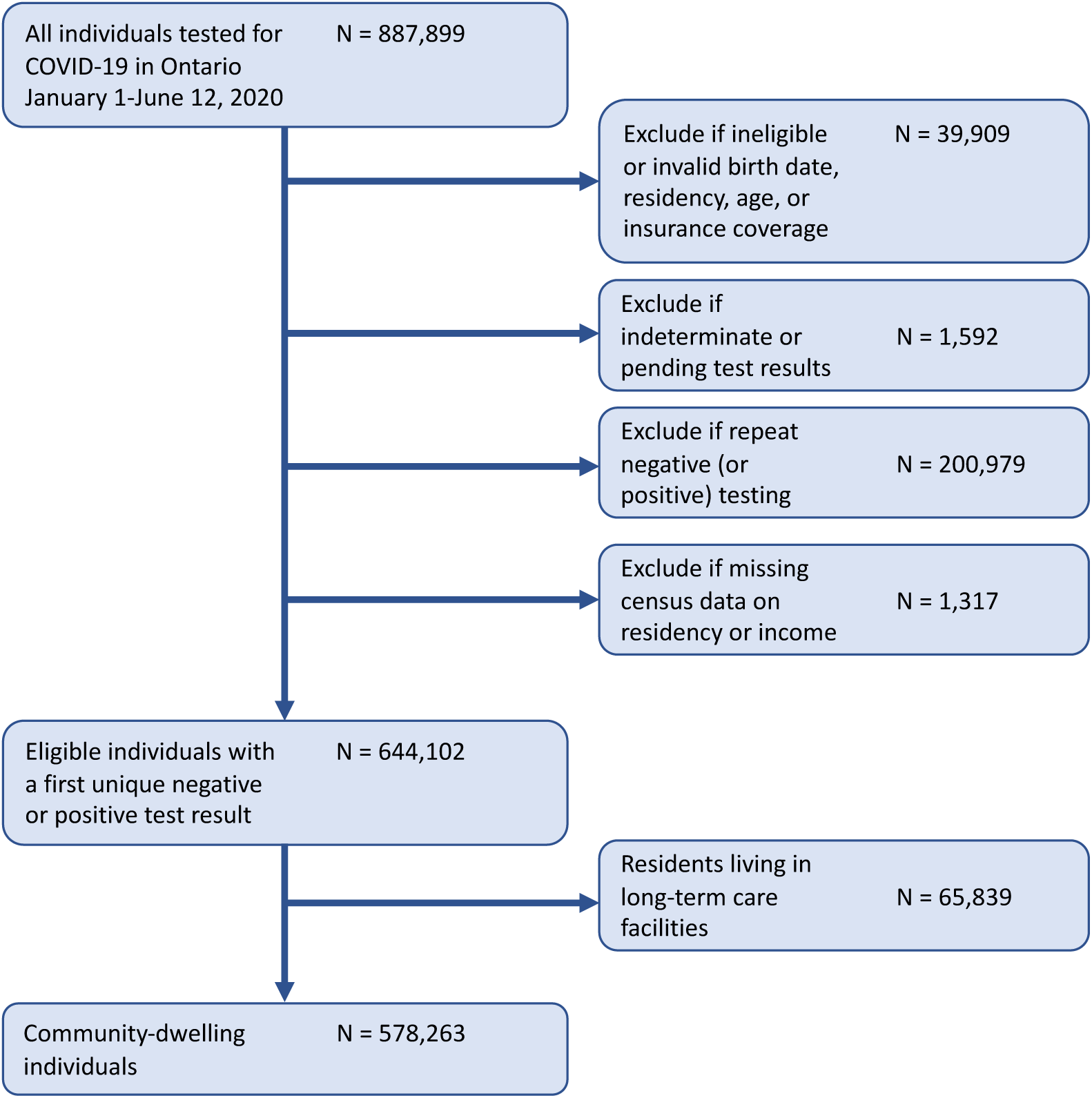
Study Eligibility Flow Chart

**Supplemental Figure 3.**
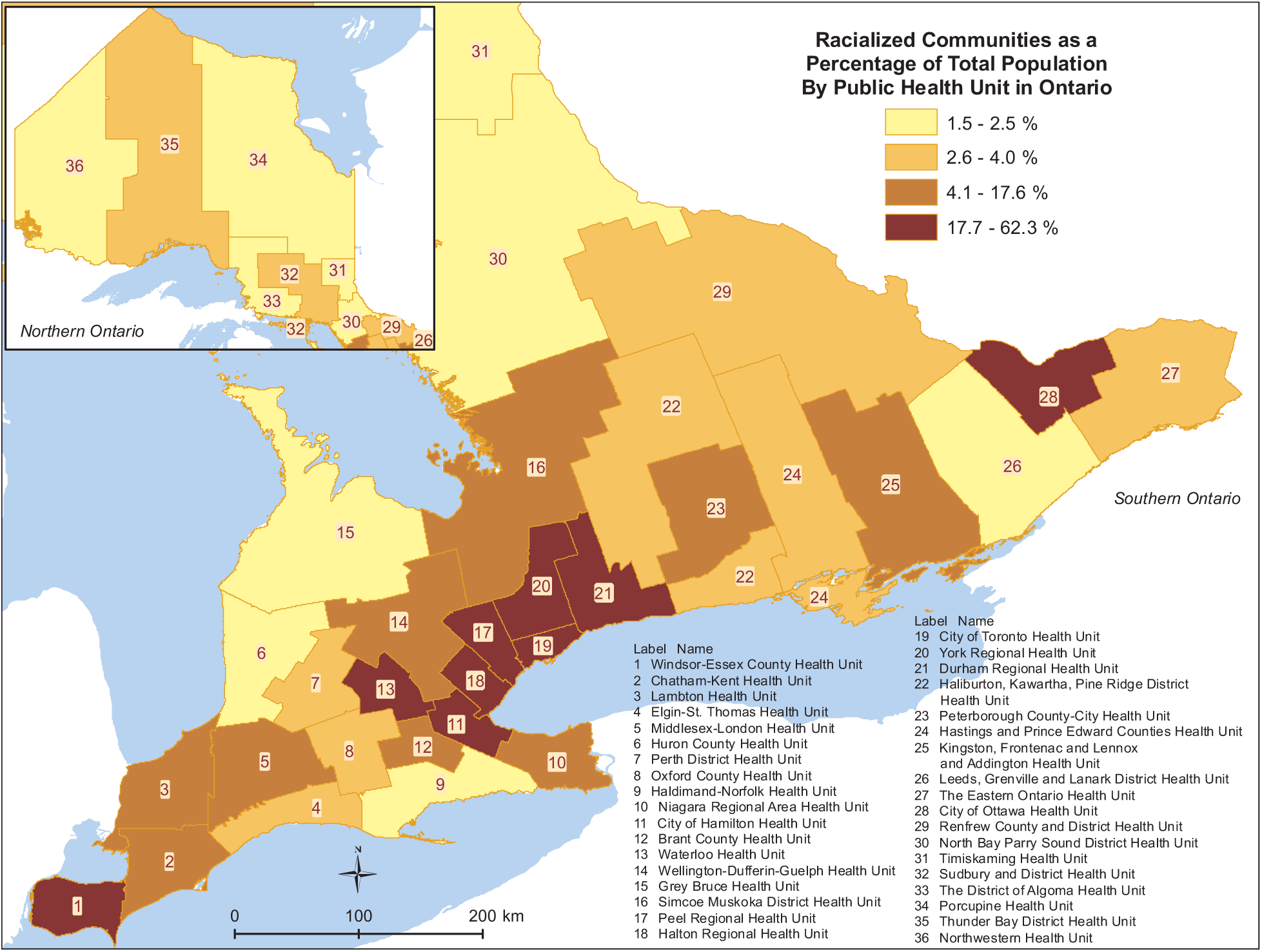
Distribution of Regional Racial/Ethnic Diversity in Ontario as a Percentage of Total Population by Public Health Unit. The percentage of racialized communities categorized by public health unit (PHU). PHUs were divided into quartiles; the median rate was 4.0% (interquartile range, 2.5-17.6%). Racial/ethnic diversity was defined as the regional proportion of individuals who self-identified as Black, South Asian, Chinese, Filipino, Latin American, Arab, Southeast Asian, West Asian, Korean and Japanese according to 2016 Census data.

**Supplemental Figure 4.**
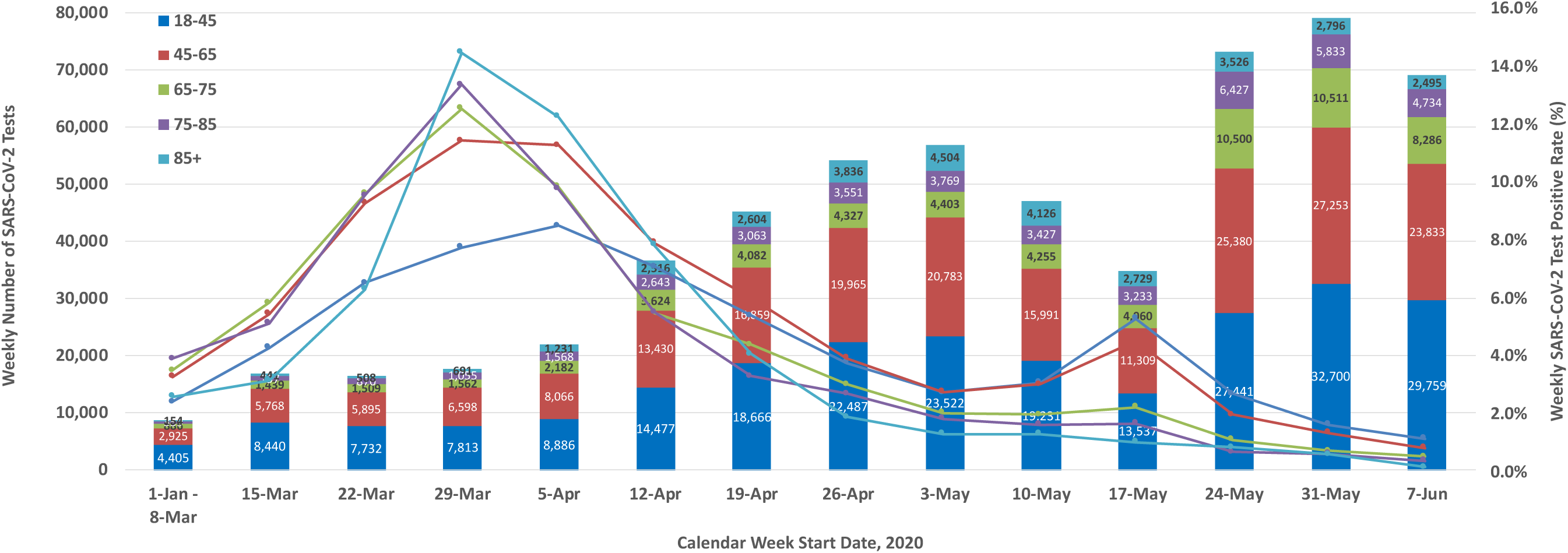
Weekly number of community-dwelling individuals tested for SARS-CoV-2, and share of tests that were positive, stratified by age group during the first wave of the pandemic in Ontario, Canada. Time represented by the start of the calendar week with the weeks of January 1 through March 8 consolidated given low initial test counts. Age groups: 18-45 years (blue); 45-65 years (red); 65-75 years (green); 75-85 years (purple); 85 years and older (light blue). The number of weekly counts attributable to an age group is represented in each stacked column (primary y-axis). The weekly positive test rate attributable to an age group is represented in each line graph (secondary y-axis). The share of positive test results among younger community-dwelling patients (18-65 years) overcoming those of older community-dwelling patients (65 years and older) during the first wave of the pandemic in Ontario occurred by the week of April 12, 2020.

**Supplemental Table 1.**
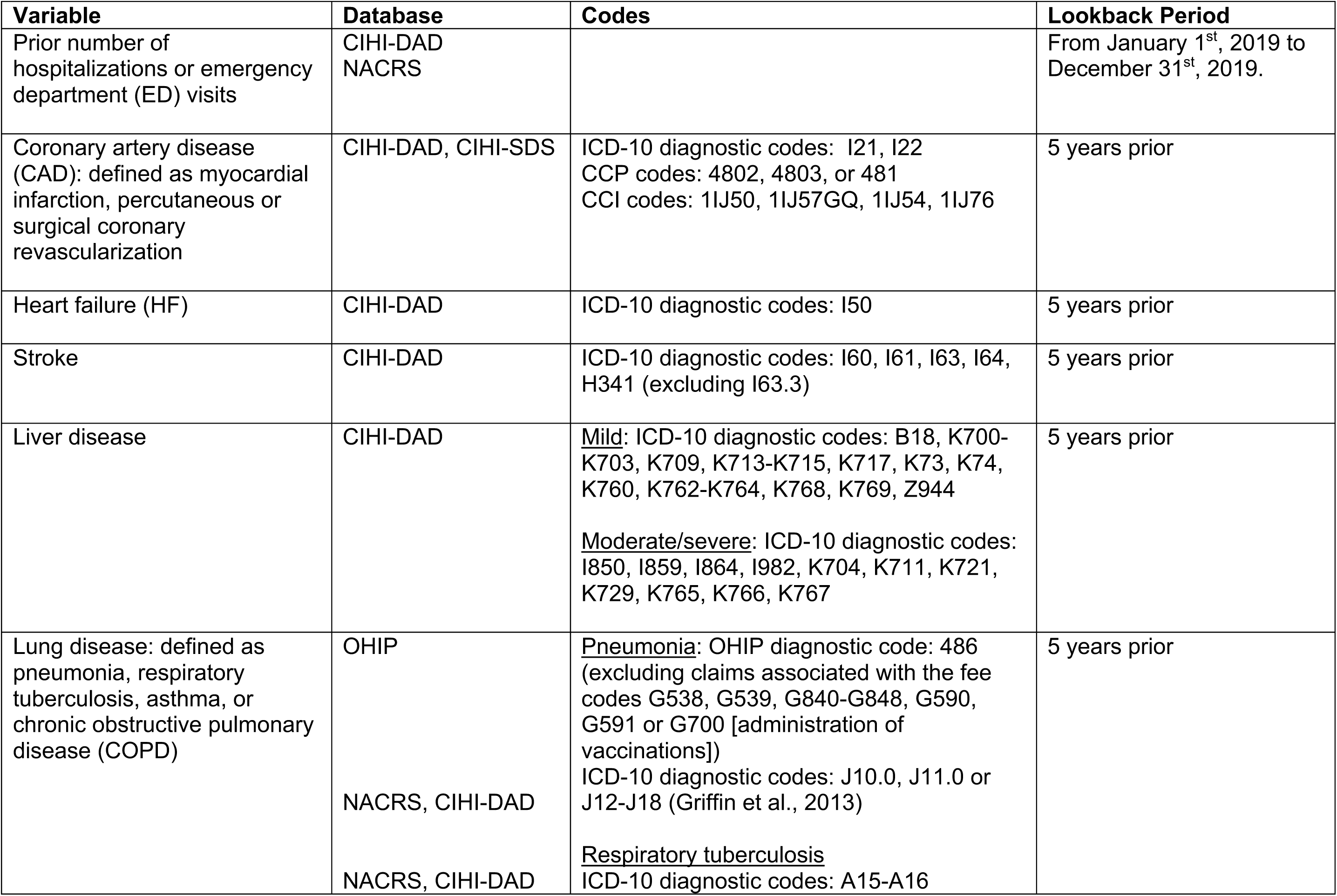
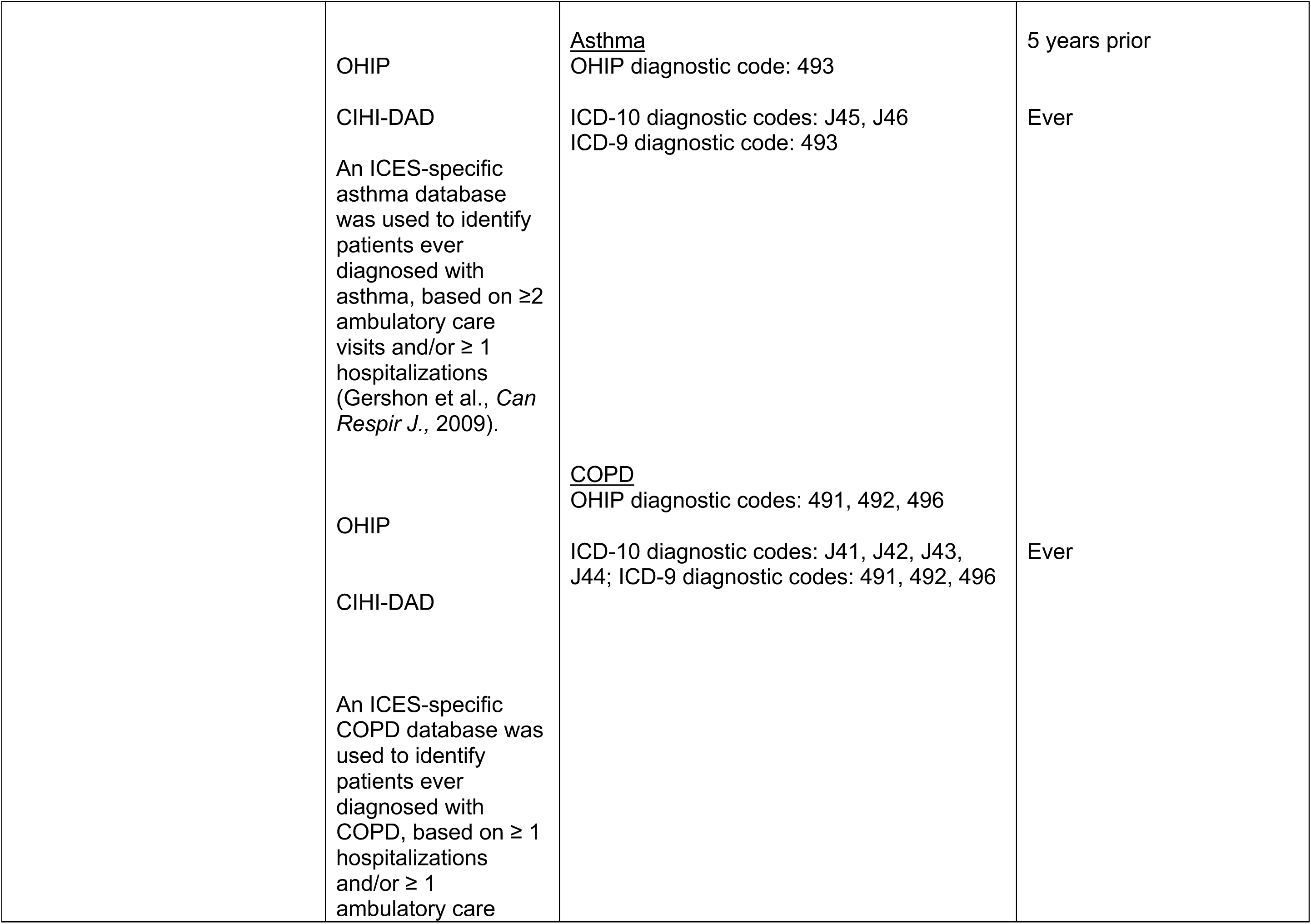
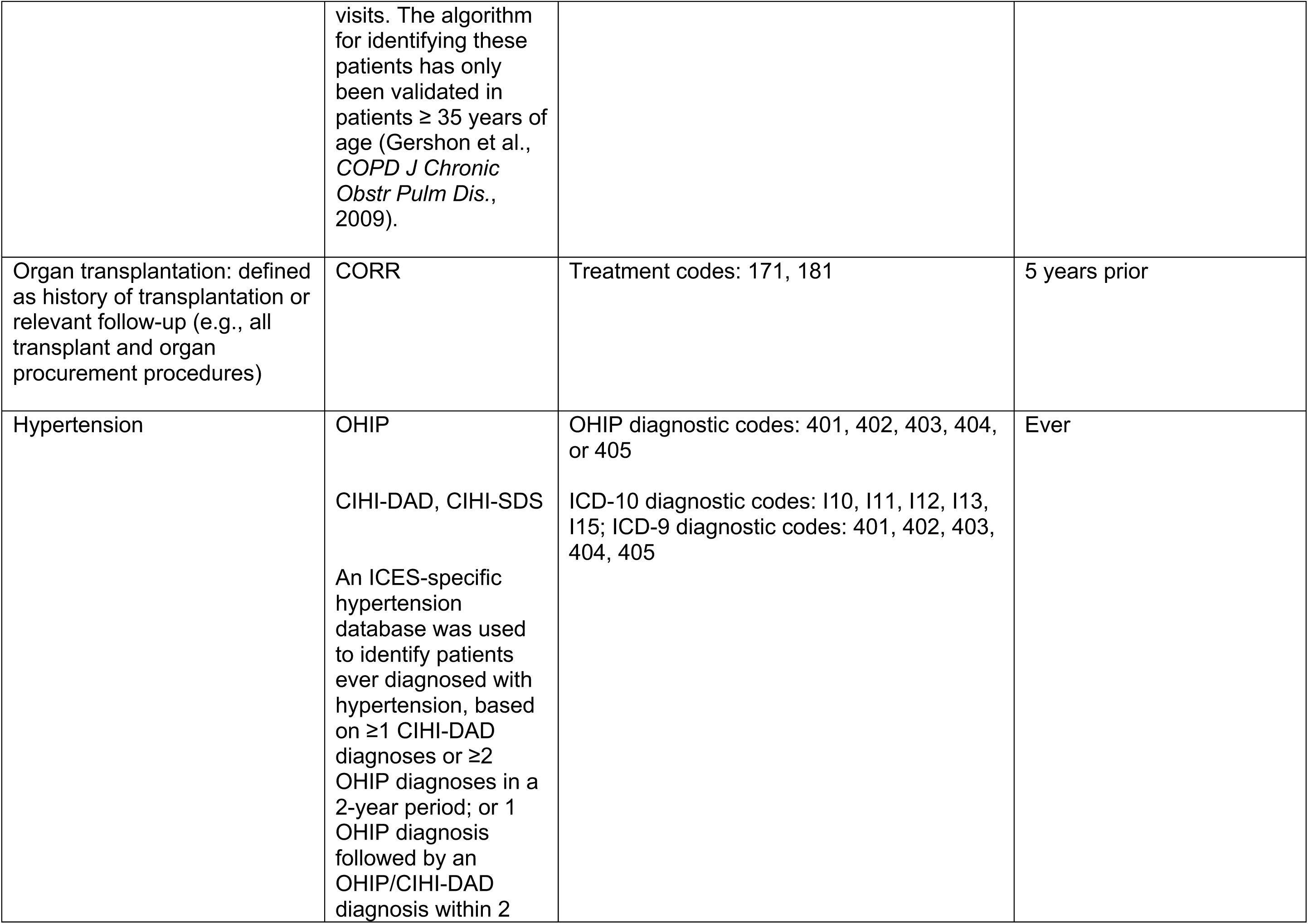
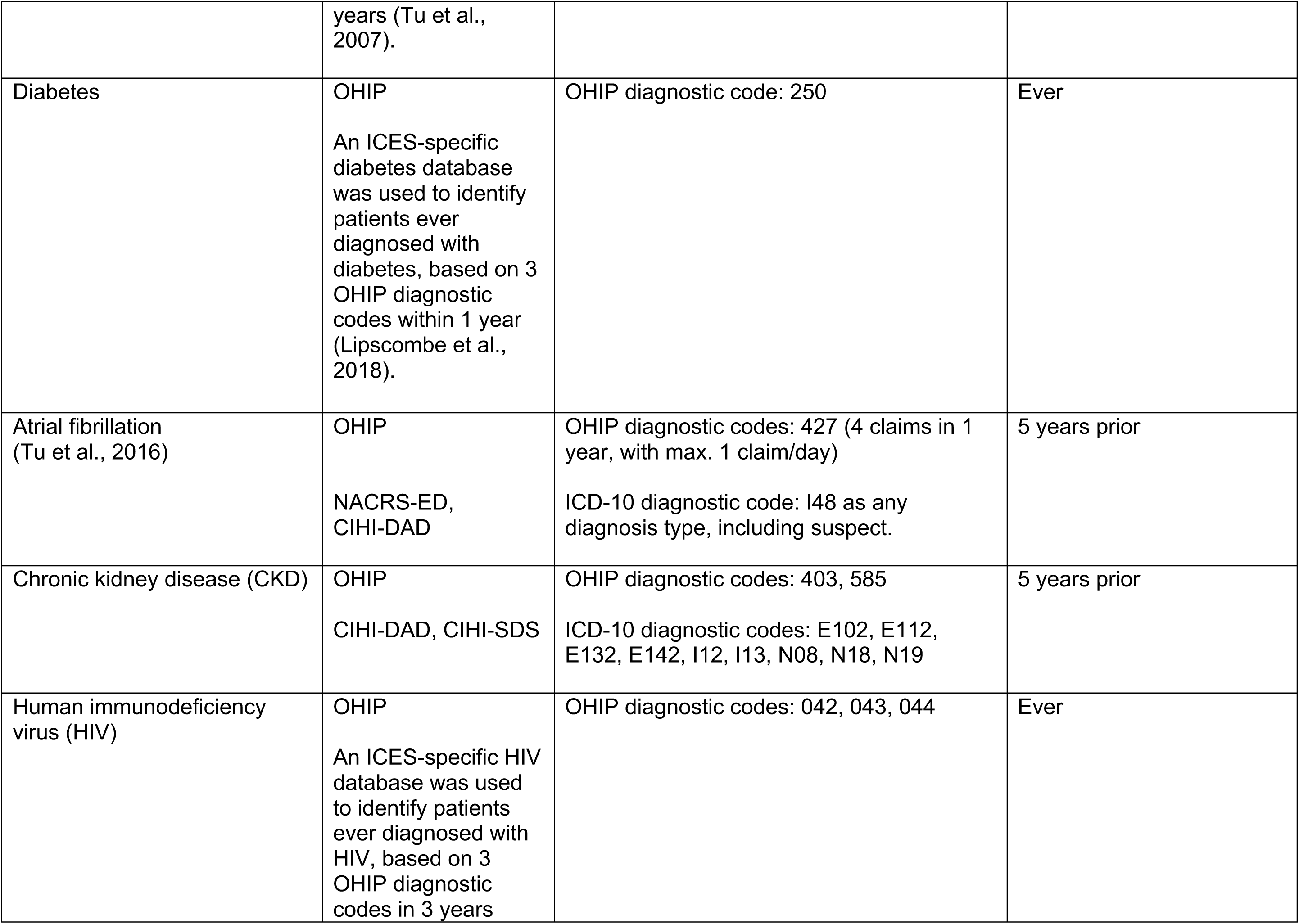
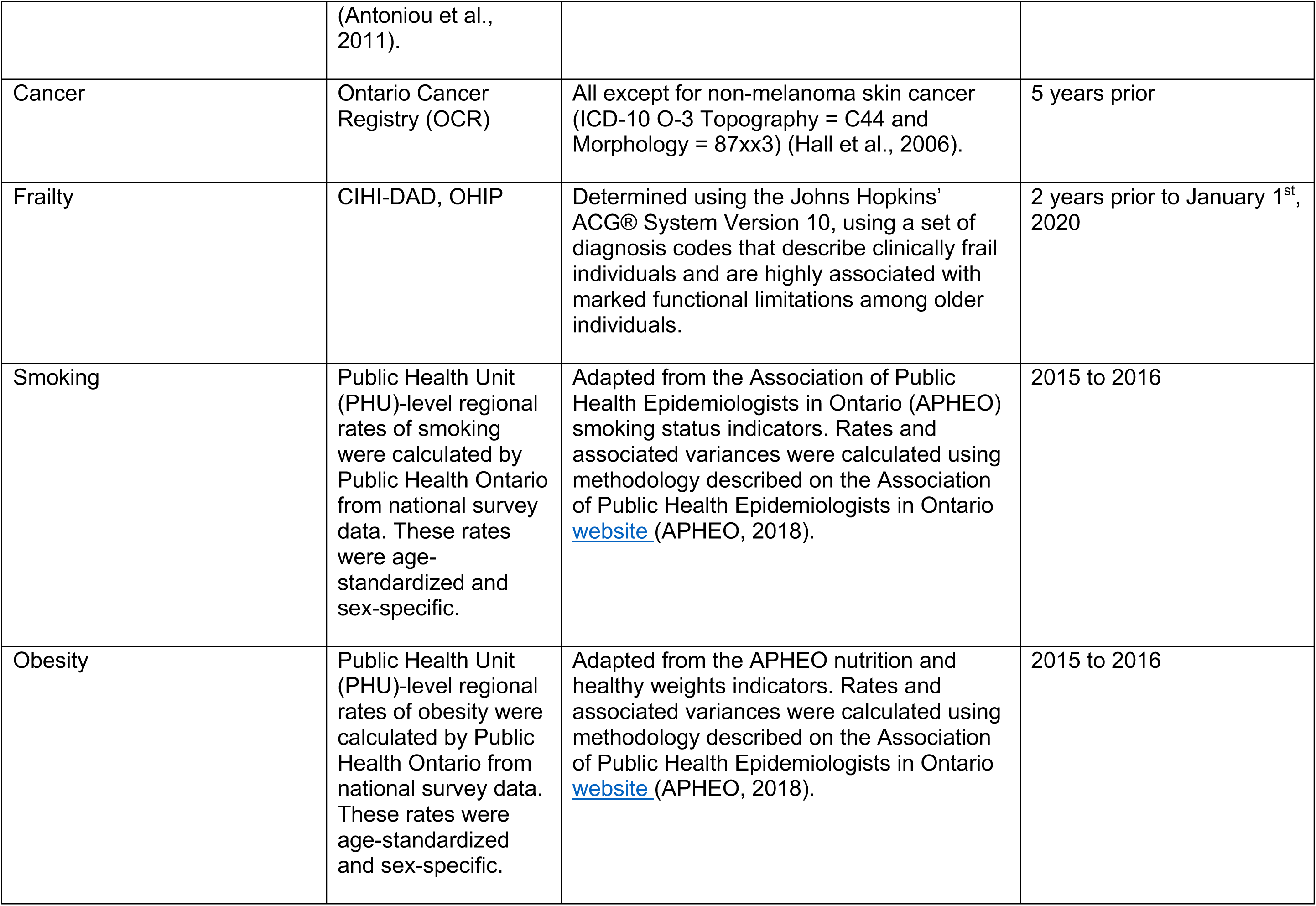
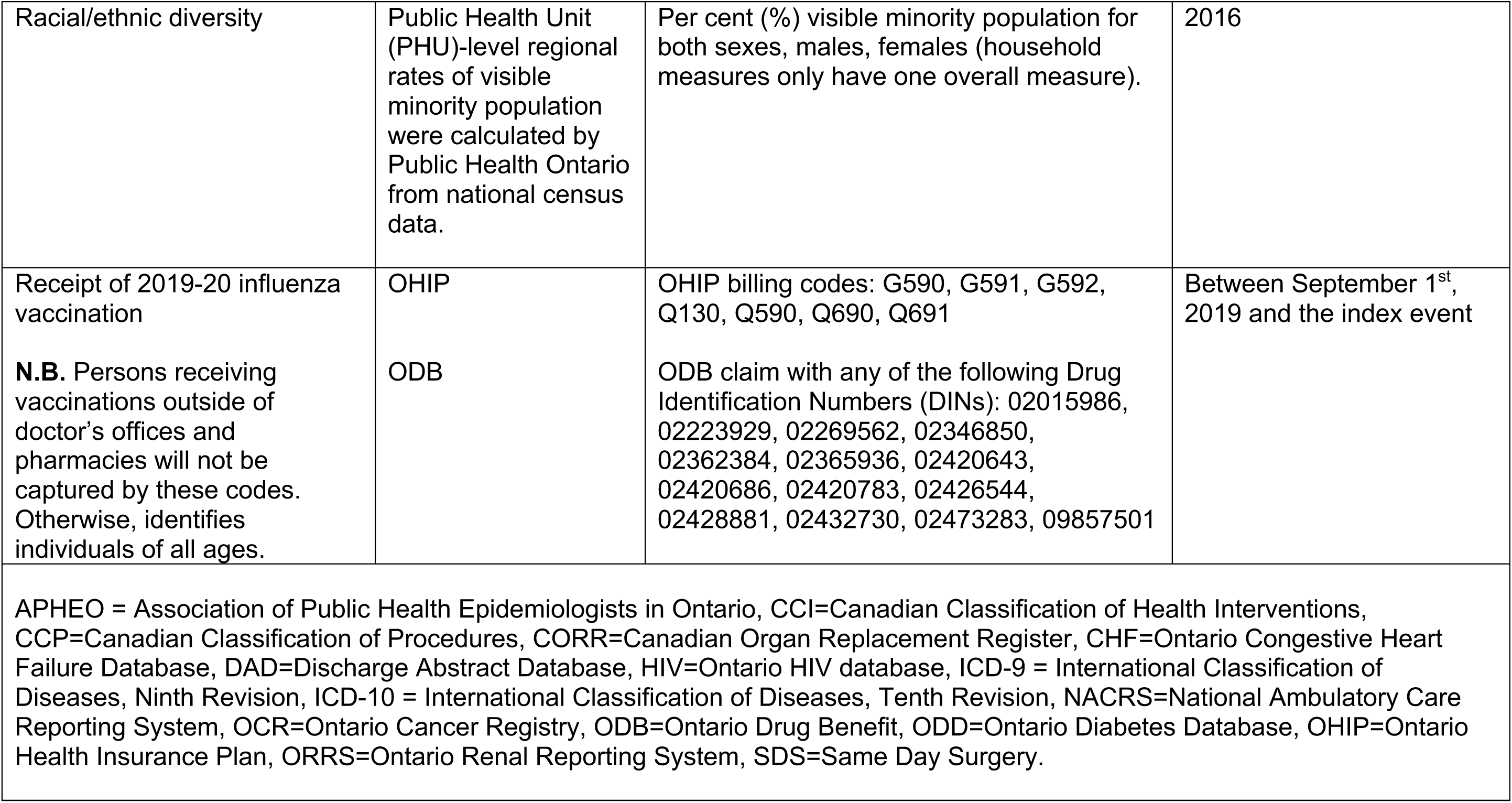
Administrative Health Data Codes Defining Baseline Clinical Diagnoses

**Supplemental Table 2.** Baseline Characteristics of Long-Term Care-Dwelling Individuals with and without SARS-CoV-2 Infection in Ontario

| Characteristic |  | Negative Test<br>(n=60,917) | Positive Test<br>(n=4,922) | SD | P-value |
| --- | --- | --- | --- | --- | --- |
| Week of testing (start date) | 0-10 (Jan. 1 – Mar. 8) | 164 (0.3%) | 16 (0.3%) | 0.01 | <0.001 |
|  | 11 (Mar. 15) | 450 (0.7%) | 36 (0.7%) | 0 |  |
|  | 12 (Mar. 22) | 582 (1.0%) | 71 (1.4%) | 0.04 |  |
|  | 13 (Mar. 29) | 1,303 (2.1%) | 411 (8.4%) | 0.28 |  |
|  | 14 (Apr. 5) | 2,244 (3.7%) | 989 (20.1%) | 0.52 |  |
|  | 15 (Apr. 12) | 6,666 (10.9%) | 1,723 (35.0%) | 0.60 |  |
|  | 16 (April 19) | 10,206 (16.8%) | 889 (18.1%) | 0.03 |  |
|  | 17 (April 26) | 12,798 (21.0%) | 321 (6.5%) | 0.43 |  |
|  | 18 (May 3) | 17,043 (28.0%) | 303 (6.2%) | 0.61 |  |
|  | 19 (May 10) | 6,412 (10.5%) | 90 (1.8%) | 0.37 |  |
|  | 20 (May 17) | 708 (1.2%) | 37 (0.8%) | 0.04 |  |
|  | 21 (May 24) | 838 (1.4%) | 22 (0.4%) | 0.10 |  |
|  | 22 (May 31) | 866 (1.4%) | 5-10 (0.2%) | 0.14 |  |
|  | 23 (June 7) | 637 (1.0%) | ≤ 5 (0.1%) | 0.13 |  |
| Age, years |  | 85 (77-91) | 85 (77-91) | 0.03 | 0.03 |
| Age group, years | 18-45 | 472 (0.8%) | 34 (0.7%) | 0.01 | 0.50 |
|  | 45-65 | 4,632 (7.6%) | 399 (8.1%) | 0.02 |  |
|  | 65-75 | 8,165 (13.4%) | 685 (13.9%) | 0.01 |  |
|  | 75-85 | 17,701 (29.1%) | 1,416 (28.8%) | 0.01 |  |
|  | ≥85 | 29,947 (49.2%) | 2,388 (48.5%) | 0.01 |  |
| Sex | Female | 41,261 (67.7%) | 3,193 (64.9%) | 0.06 | <0.001 |
|  | Male | 19,656 (32.3%) | 1,729 (35.1%) | 0.06 |  |
| Income quintile | 1 | 18,162 (29.8%) | 1,501 (30.5%) | 0.01 | <0.001 |
|  | 2 | 13,087 (21.5%) | 1,313 (26.7%) | 0.12 |  |
|  | 3 | 11,099 (18.2%) | 903 (18.3%) | 0 |  |
|  | 4 | 10,126 (16.6%) | 558 (11.3%) | 0.15 |  |
|  | 5 | 8,443 (13.9%) | 647 (13.1%) | 0.02 |  |
| Community size | <10,000 | 8,307 (13.6%) | 162 (3.3%) | 0.38 | <0.001 |
|  | 10,000-100,000 | 7,872 (12.9%) | 219 (4.4%) | 0.30 |  |
|  | 100,000-500,000 | 16,005 (26.3%) | 441 (9.0%) | 0.47 |  |
|  | 500,000-1.5 million | 10,146 (16.7%) | 640 (13.0%) | 0.10 |  |
|  | ≥1.5 million | 18,587 (30.5%) | 3,460 (70.3%) | 0.87 |  |
| Canadian immigrant |  | 3,509 (5.8%) | 476 (9.7%) | 0.15 | <0.001 |
| Regional smoking rate, % |  | 13 (9-17) | 11 (9-12) | 0.68 | <0.001 |
| Regional obesity rate, % |  | 22 (15-25) | 15 (15-21) | 0.65 | <0.001 |
| Regional racial diversity rate, % |  | 18 (4-50) | 51 (27-52) | 0.91 | <0.001 |

Supplemental Table 2 Continued. Baseline Characteristics of Long-Term Care-Dwelling Individuals with and without SARS-CoV-2 Infection in Ontario
| Characteristic |  | Negative Test<br>(n=557,739) | Positive Test<br>(n=20,524) | SD | P-value |
| --- | --- | --- | --- | --- | --- |
| **Clinical risk factors** |  |  |  |  |  |
| Hypertension |  | 48,500 (79.6%) | 4,003 (81.3%) | 0.04 | 0.004 |
| Hypertension duration, years |  | 19 (13-25) | 19 (12-25) | 0 | 0.98 |
| Diabetes |  | 17,192 (28.2%) | 1,592 (32.3%) | 0.09 | <.001 |
| Coronary artery disease |  | 6,688 (11.0%) | 483 (9.8%) | 0.04 | 0.012 |
| Heart failure |  | 5,836 (9.6%) | 515 (10.5%) | 0.03 | 0.044 |
| Stroke |  | 4,193 (6.9%) | 397 (8.1%) | 0.04 | 0.002 |
| Atrial fibrillation |  | 9,709 (15.9%) | 897 (18.2%) | 0.06 | <.001 |
| Chronic kidney disease |  | 8,618 (14.1%) | 839 (17.0%) | 0.08 | <.001 |
| HIV |  | 80 (0.1%) | 10 (0.2%) | 0.02 | 0.189 |
| Organ transplantation |  | 71 (0.1%) | 8 (0.2%) | 0.01 | 0.37 |
| Cancer |  | 2,340 (3.8%) | 182 (3.7%) | 0.01 | 0.614 |
| Liver disease |  | 630 (1.0%) | 60 (1.2%) | 0.02 | 0.221 |
| Lung disease |  | 24,575 (40.3%) | 2,079 (42.2%) | 0.04 | 0.009 |
| Hospitalization or ED visits in 2019 | 0 | 19,985 (32.8%) | 1,482 (30.1%) | 0.06 | <0.001 |
|  | 1-2 | 22,089 (36.3%) | 1,885 (38.3%) | 0.04 |  |
|  | 3+ | 18,843 (30.9%) | 1,555 (31.6%) | 0.01 |  |
| Frailty |  | 45,831 (75.2%) | 3,810 (77.4%) | 0.05 | <0.001 |
| Influenza vaccination in 2019 |  | 10,508 (17.2%) | 563 (11.4%) | 0.17 | <0.001 |

Continuous variables are reported as median (interquartile range). Abbreviations: ED: emergency department; HIV: human immunodeficiency virus; SD: standardized difference.

**Supplemental Table 3.** Predictors of SARS-CoV-2 Infection among Long-Term Care-Dwelling Individuals in Ontario, Canada.

| Characteristic |  | OR (95% CI) | P-value |
| --- | --- | --- | --- |
| Week of testing (start date) | 0-10 (Jan. 1 – Mar. 8) | Referent | <0.0001 |
|  | 11 (Mar. 15) | 0.94 (0.50-1.75) |  |
|  | 12 (Mar. 22) | 1.60 (0.89-2.86) |  |
|  | 13 (Mar. 29) | 4.52 (2.64-7.73) |  |
|  | 14 (Apr. 5) | 6.29 (3.70-10.68) |  |
|  | 15 (Apr. 12) | 2.84 (1.68-4.80) |  |
|  | 16 (April 19) | 1.18 (0.70-2.00) |  |
|  | 17 (April 26) | 0.41 (0.24-0.70) |  |
|  | 18 (May 3) | 0.28 (0.16-0.48) |  |
|  | 19 (May 10) | 0.23 (0.13-0.40) |  |
|  | 20 (May 17) | 0.64 (0.35-1.20) |  |
|  | 21 (May 24) | 0.30 (0.15-0.58) |  |
|  | 22 (May 31) | 0.11 (0.05-0.26) |  |
|  | 23 (June 7) | 0.09 (0.03-0.24) |  |
| Age group, years | 18-45 | Referent | 0.75 |
|  | 45-65 | 1.14 (0.77-1.69) |  |
|  | 65-75 | 1.11 (0.75-1.64) |  |
|  | 75-85 | 1.08 (0.73-1.60) |  |
|  | ≥85 | 1.06 (0.72-1.56) |  |
| Sex | Female | Referent |  |
|  | Male | 1.07 (0.92-1.25) | 0.37 |
| Rural dwelling |  | 0.79 (0.65-0.97) | 0.02 |
| Income quintile | 1 | 0.90 (0.81-1.01) |  |
|  | 2 | 1.11 (0.99-1.24) |  |
|  | 3 | 1.01 (0.90-1.14) |  |
|  | 4 | 0.75 (0.66-0.86) |  |
|  | 5 | Referent | <0.0001 |
| Canadian immigrant |  | 0.98 (0.87-1.09) | 0.69 |
| Frailty |  | 0.91 (0.84-0.98) | 0.02 |
| Hospitalization or ED visits in 2019 | 0 | Referent | 0.047 |
|  | 1-2 | 1.10 (1.02-1.20) |  |
|  | 3+ | 1.08 (0.99-1.18) |  |
| Hypertension |  | 1.00 (0.92-1.10) | 0.96 |
| Diabetes |  | 1.12 (1.04-1.20) | 0.004 |
| Coronary artery disease |  | 0.87 (0.78-0.98) | 0.26 |
| Heart failure |  | 0.94 (0.83-1.05) | 0.26 |
| Stroke |  | 1.10 (0.98-1.25) | 0.12 |
| Atrial fibrillation |  | 1.07 (0.98-1.17) | 0.14 |
| Chronic kidney disease |  | 0.94 (0.85-1.03) | 0.17 |
| HIV |  | 0.69 (0.34-1.41) | 0.31 |
| Organ transplantation |  | 0.72 (0.32-1.60) | 0.42 |
| Cancer |  | 0.93 (0.68-1.23) | 0.37 |
| Liver disease |  | 0.92 (0.68-1.23) | 0.57 |
| Lung disease |  | 0.94 (0.88-1.01) | 0.08 |
| Influenza vaccination in 2019 |  | 0.75 (0.67-0.83) | <0.0001 |
| Regional smoking rate, per 1% |  | 1.02 (0.99-1.06) | 0.20 |
| Regional obesity rate, per 1% |  | 0.99 (0.97-1.00) | 0.12 |
| Regional racial diversity rate, per 1% |  | 1.05 (1.03-1.08) | <0.0001 |

Odds ratios were determined from a multivariable logistic regression model that incorporated PHU-specific random effects. For continuous variables, results are per 1-unit increase in rate. Abbreviations: CI: confidence intervals; ED: emergency department; HIV: human immunodeficiency virus; OR; odds ratio.

## Notes

### Competing Interest Statement

The authors have declared no competing interest.

### Author Declarations

ICES has obtained ethical approval (and repeats this review tri-annually) for its privacy and security policies, procedures, and practices. Each research project that is conducted at ICES is also subject to internal ethical review by the ICES Privacy and Compliance Office. Please find attached to this submission a letter with more information regarding the ethical review and approval process for this research; please do not hesitate to contact me with any questions. ICES is a prescribed entity under section 45 of Ontario's Personal Health Information Protection Act (PHIPA). Section 45 is the provision that enables analysis and compilation of statistical information related to the management, evaluation, and monitoring of, allocation of resources to, and planning for the health system. Section 45 authorizes health information custodians to disclose personal health information to a prescribed entity, like ICES, without consent for such purposes. Projects conducted wholly under section 45, by definition, do not require review by a Research Ethics Board. As a prescribed entity, ICES must submit to trio-annual review and approval of its privacy and security policies, procedures and practices by Ontario's Information and Privacy Commissioner. These include policies, practices and procedures that require internal review and approval of every project by ICES' Privacy and Compliance Office. ICES was approved by the Commissioner for a fifth time in 2017.

